# Replicating clinical placebo effects in computational trials: Bridging the gap between in silico and clinical studies

**DOI:** 10.1101/2024.10.18.24315721

**Authors:** Paula Dominguez-Gomez, Constantine Butakoff, Georg Rast, Borje Darpo, Mariano Vazquez, Jazmin Aguado-Sierra

**Author notes:** Corresponding author at: Elem Biotech, Via Laietana, 26, 4th Floor B, Barcelona 08003, Spain. Email address:* (Paula Dominguez-Gomez).

## Abstract

This study replicates human cardiac safety clinical trials using computational models, which include diurnal hormonal placebo effects, aiming to create fast and efficient drug screening tools to assess QT interval prolongation preclinically. A virtual cardiac population that simulates sex-specific electrophysiological variability influenced by diurnal hormonal cycles was created to closely mirror human physiology. The goal is to assess the impact of these hormone dynamics on placebo-induced responses and evaluate the virtual population’s accuracy in reflecting clinical trial outcomes.

The study showed that the virtual population model successfully replicated about 50% of the observed QT variability seen in placebo groups. The computational framework facilitated direct comparison with human clinical trials, achieving critical concentrations, i.e., concentrations that cause a mean QTc effect of 10 ms, within a 0.6-fold range of clinical results. Additionally, the exposure-response slopes exhibited relative errors averaging 43%, which translates into mean absolute errors of 6-7 ms in comparison to clinical data. These results highlight its potential to reproduce human physiology and provide results consistent with real-world human clinical trials. Additionally, a comparison with a previously published statistical placebo model further supports this approach. By approximating the diurnal hormonal variations, as a component of the mechanistic basis of placebo, this method offers a robust decision-making tool for early-stage drug safety assessment.

**Highlights:**

- Diurnal hormonal fluctuations significantly contribute to intersubject variability observed in placebo responses.
- Modeling conservative diurnal hormonal variations can reproduce approximately 50% of QT interval prolongation variability within placebo groups.
- Comprehensive human clinical trials for cardiac safety drug testing, including placebo effects and placebo-corrected QT interval prolongation, can be accurately replicated through virtual human computational models showing mean absolute errors of 6-7 ms.

**Graphical Abstract:** 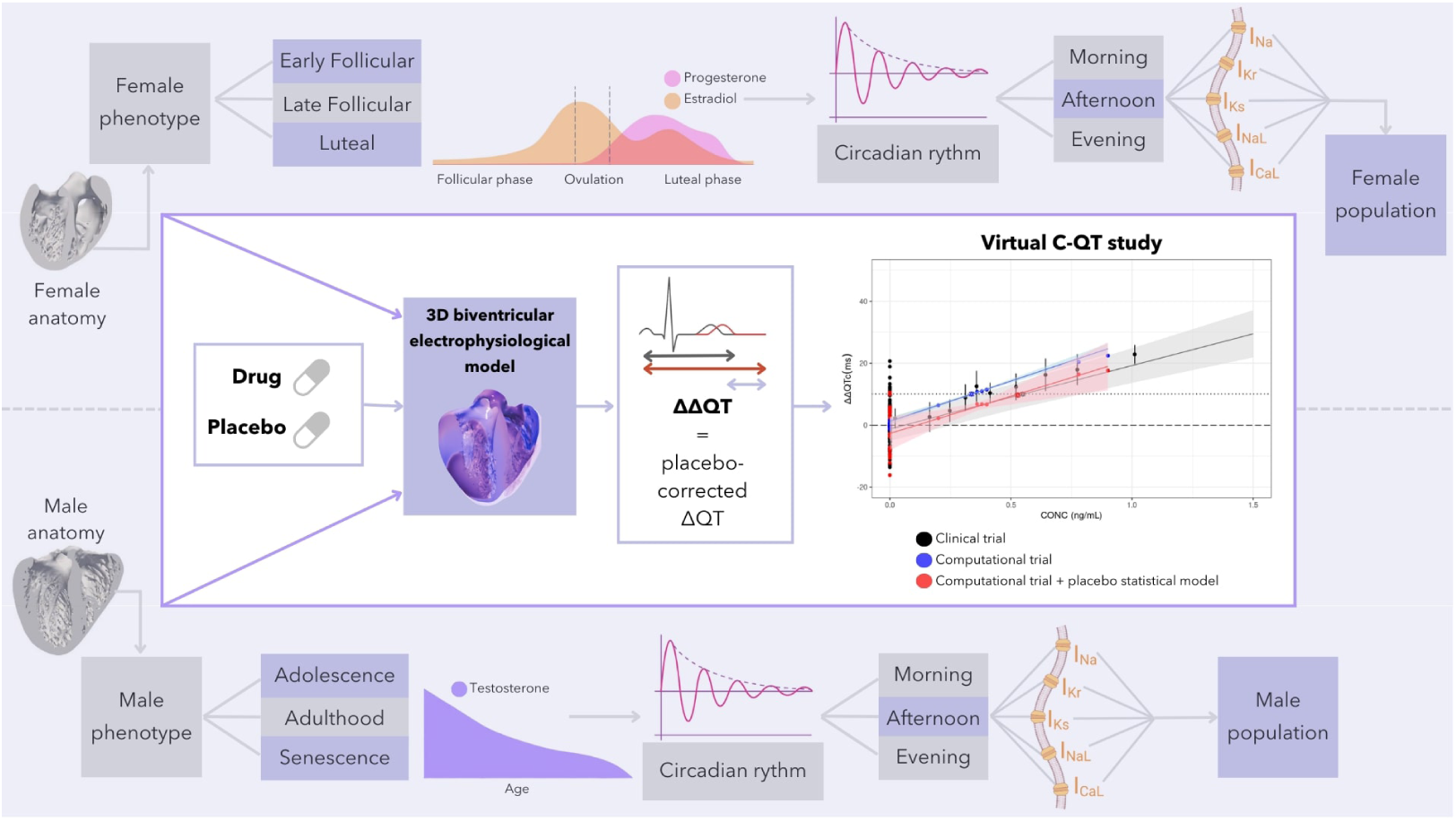

## 1. Introduction

In recent decades, several pharmaceutical compounds have been cancelled during clinical development or withdrawn from the market due to their potential to induce lethal arrhythmias. These arrhythmias are often linked to the drug’s effects on cardiac electrophysiology, through blocking cardiac ion channels (Mirams et al., 2011a). A critical marker of this risk is the QTc interval, measured on an electrocardiogram (ECG), which reflects the time for the heart’s ventricles to depolarize and repolarize. Prolongation of the QTc interval (ΔQTc) can lead to torsades de pointes, a dangerous form of arrhythmia. To assess this risk, regulatory agencies like the U.S. Food and Drug Administration (FDA) and the European Medicines Agency (EMA) mandate thorough QT (TQT) studies, alternatively, early clinical studies sufficiently powered to exclude a small effect on the QTc interval before proceeding into late stage clinical trials.

Placebo-controlled clinical trials are essential in this process, as they help isolate the effects of the drug by accounting for variability due to factors unrelated to the treatment. This variability, termed the placebo effect, arises from a range of influences, including intersubject variability and intrasubject changes such as circadian rhythms, environmental shifts, meals, or stress. In clinical trials, placebo-corrected ΔQT (ΔΔQTc) is computed to correct for such time-dependent drifts in baseline QT intervals (Garnett et al., 2018). One of the main mechanistic contributors to baseline QTc variability is sex steroid hormonal fluctuation, which differs between sexes. For example, in females, QTc intervals vary across the menstrual cycle, being longer in the follicular phase and shorter in the luteal phase due to changes in estrogen and progesterone levels (Furukawa and Kurokawa, 2007; James et al., 2007). Progesterone shortens QTc, while estrogen lengthens it (Nakamura et al., 2007; Kurokawa et al., 2008). In males, testosterone influences QTc, with reductions during puberty and increases with age-related testosterone decline (Bidoggia et al., 2000; Bai et al., 2005). While these higher magnitude fluctuations occur over weeks or years, diurnal fluctuations, such as those induced by circadian rhythms, can affect QTc on a much shorter time scale and may more directly confound the QT effects of drugs with typical clearance rates.

Despite that the state of the art are human clinical trials, there is growing interest in using computational models to assess proarrhythmic risk, as these models can reduce the time and costs associated with drug development. Computational models have shown promise in predicting drug effects on cardiac electrophysiology by simulating how compounds interact with cardiac ion channels. Initiatives like the Comprehensive In Vitro Proarrhythmia Assay (CiPA) aim to develop mechanistic-based assessments that go beyond the traditional focus on hERG channel block and QTc prolongation (Park et al., 2019). While many existing computational models focus on single-cell or population-level simulations (Mirams et al., 2011b; Yang and Clancy, 2012; Dutta et al., 2017; Trovato et al., 2022; Passini et al., 2021), fewer studies employ more comprehensive multiscale models that integrate biventricular anatomies, which have the potential to capture complex drug-induced proarrhythmic mechanisms (Okada et al., 2018; Hwang et al., 2019; Margara et al., 2021). Crucially, current computational models do not account for the placebo effect, a key element in clinical trials.

This work addresses this gap by extending the capabilities of current mechanistic models to simulate a full clinical trial, including both treatment and placebo groups. Building on the approach developed by Aguado-Sierra et al. (2024), the aim is to mechanistically reproduce the placebo effect in a virtual population. It is hypothesized that diurnal hormonal variations are a primary driver of placebo-induced QTc variability, and the model incorporates sex differences, ion channel density variability, and hormonal fluctuations to simulate a normal virtual human population. This allows to compute virtual ECGs for each subject, quantify ΔQTc for both control and treatment groups, and calculate ΔΔQTc as done in clinical trials.

To validate the model, clinical trial data from two drugs, moxifloxacin and dofetilide (Darpo et al., 2015), were used and the results were compared with the model published by Minocha et al. (2019). This approach enabled an assessment of the degree to which the assumptions align with the total placebo effect observed in clinical practice, providing a more comprehensive framework for cardiac safety assessments in drug development.

## 2. Methods

This in silico study was performed using the same methodology published in Aguado-Sierra et al. (2022, 2024) for the generation of a virtual human population using high resolution human biventricular anatomies solved using finite elements using Alya (Vázquez et al., 2016; Santiago et al., 2018), a multi-physics, multi-scale and finite element-based simulation code developed at the Barcelona Supercomputing Center and commercialized by ELEM Biotech.

### 2.1. Anatomical Data

For the current study, one female and one male biventricular heart anatomies reconstructed from high-resolution magnetic resonance imaging collected from the University of Minnesota’s Visible Heart Lab library (Visible Heart Laboratories, 2021) were included. The detailed biventricular anatomy of both the female (20yo, BMI 19.1) and male (39yo, BMI 25.1) ex-vivo human hearts were segmented from high-resolution magnetic resonance imaging (MRI) data. These hearts were cannulated and perfusion-fixed under a pressure of 40-50 mmHg, with 10% phosphate buffered formalin, which preserved them at end-diastolic state. High resolution images of these specimens were acquired via a 3T Siemens scanner: with 0.44×0.44 mm in-plane resolution and slice thickness of 1 to 1.7 mm, which provided detailed endocardial trabeculae and false tendons with approximately 1 mm^2^ cross sectional area. The measured myocardial volume of the heart with anatomy ID 311 (female, Figure 1b) is 170.6 cm^3^, and its trabecular volume is 8.7%. The measured myocardial volume of the heart with anatomy ID 132 (male, Figure 1a) is 394.2 cm^3^, and its trabecular volume is 16.3%. A detailed description of both anatomies can be found in Gonzalez-Martin et al. (2023).

**Figure 1:**
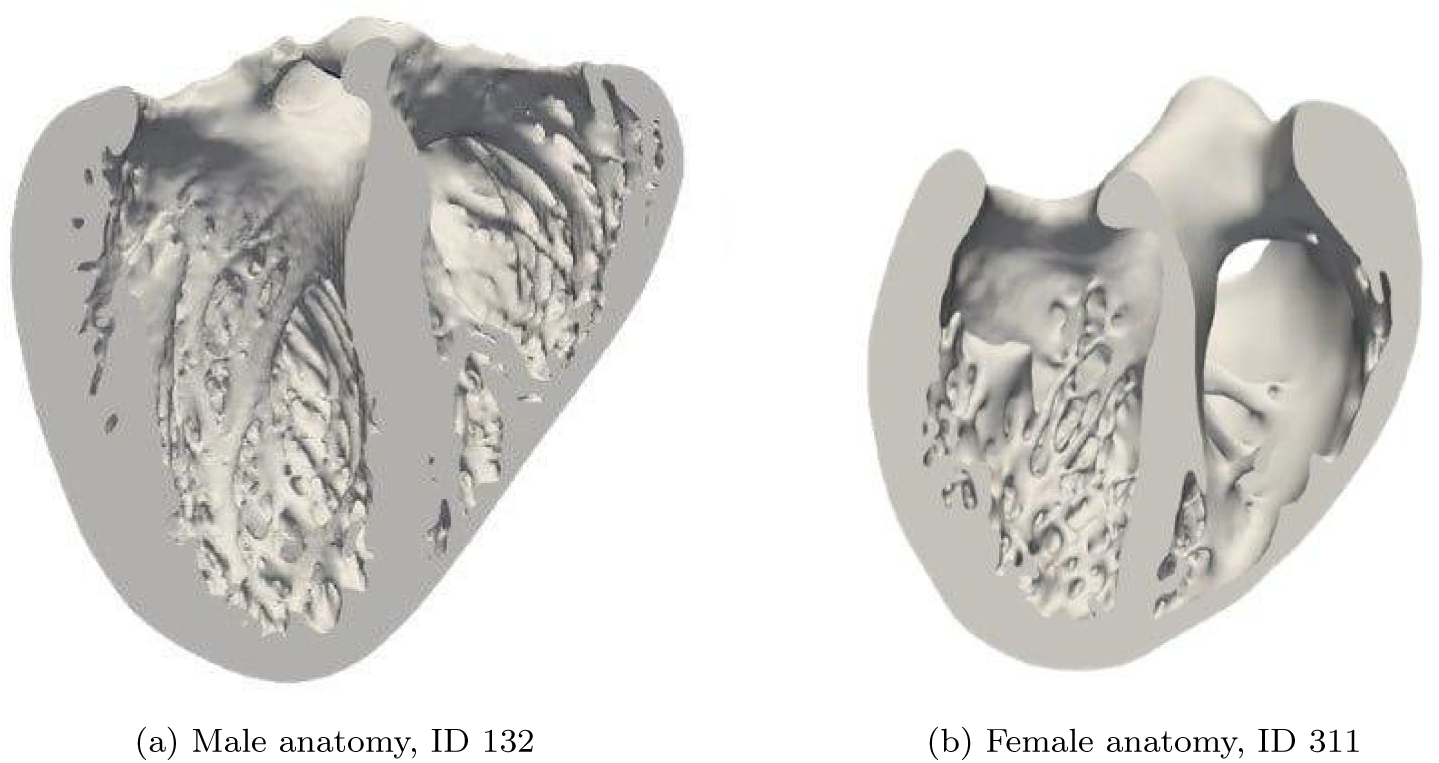
Detailed biventricular anatomies employed.

### 2.2. Mesh Construction and Tissue Properties

Volumetric finite element meshes (37.3 million tetrahedral elements for heart ID 311 and 86.3 million tetrahedral elements for heart ID 132) were created using ANSA^1^ (Sacco, 2019), with a regular element side length of 328 *µ*m and a rule-based fiber model (Doste et al., 2019).

Transmural cell heterogeneity (endocardial, M-cells and epicardial) was assigned. The anisotropic diffusion coefficients employed to reproduce clinically observed total activation times and diffusion coefficients of the myocardium are 5.8 · 10^−3^ cm^2^/ms in the fiber direction and 1.9 · 10^−3^ cm^2^/ms in the transverse direction. The fast endocardial activation due to the Purkinje network was approximated by defining a fast endocardial layer with a thickness of 400 *µ*m. This fast conducting layer was assigned a diffusion coefficient of 1.74 · 10^−2^ cm^2^/ms in the fiber direction and 5.8 · 10^−3^ cm^2^/ms in the transverse direction. The transmural electrophysiological heterogeneity description is the same as described in Gonzalez-Martin et al. (2023).

### 2.3. Pacing Protocol

The propagation of the electrical activation was simulated by pacing the locations within the biventricular cavities for both anatomies at specific locations, as suggested by Durrer et al. (1970), as previously described in Gonzalez-Martin et al. (2023) and Aguado-Sierra et al. (2022, 2024).

### 2.4. Virtual Human Population Generation

#### 2.4.1. Electrophysiological Model

The electrophysiology model setup was done as described in Gonzalez-Martin et al. (2023) and Aguado-Sierra et al. (2024). For each heart anatomy, 32 virtual human subjects were created to account for a total population of 64 individuals, 50% of which are female and 50% male.

To create a sex-specific midmyocyte cell, given the lack of experimental data in those cells, midmyocardial ion channel conductances were defined as the mean values between the endocardial and epicardial cells (Table 1). Furthermore, apex-to-base conductance heterogeneity was incorporated to mimic the electrophysiological gradient existent between apex and base of the heart, which contributes to produce a more physiological pseudo-electrocardiogram (pseudo-ECG) signal.

**Table 1:** Sex-specific baseline conductance differences (Yang and Clancy, 2012).

| Ion Channel | Endo |  | Mid |  | Epi |  |
| --- | --- | --- | --- | --- | --- | --- |
|  | Male | Female | Male | Female | Male | Female |
| **IKs** | 1.87 | 1.5521 | 1.9074 | 1.5895 | 1.9448 | 1.6269 |
| **IKr** | 1.013 | 0.80027 | 1.058585 | 0.8433225 | 1.10417 | 0.886375 |
| **IK1** | 1.698 | 1.46028 | 1.68102 | 1.3584 | 1.66404 | 1.25652 |
| **Ito** | 1 | 0.64 | 0.8 | 0.45 | 0.6 | 0.26 |
| **INaK** | 1 | 1 | 0.97 | 0.97 | 0.94 | 0.94 |
| **IpCa** | 1.007 | 1.6112 | 0.94658 | 1.6112 | 0.88616 | 1.6112 |
| **Iup** | 1 | 1 | 1.21 | 1.21 | 1.42 | 1.42 |
| **Calmodulin** | 1 | 1.21 | 1.035 | 1.31 | 1.07 | 1.41 |
| **INaCa** | 1 | 1.15 | 1 | 1.15 | 1 | 1.15 |
| **INaL** | 2.661 | 2.661 | 2.661 | 2.661 | 2.661 | 2.661 |

Five ion channels that have the highest influence on action potential durations were selected: INa, IKr, IKs, ICaL and INaL to be modified in a combinatorial manner using the values observed in Table 2. The same percentage of variation was applied to both males and females.

**Table 2:** Scaling factors by which each ion channel is modified in a combinatorial manner to create virtual subjects with distinct electrophysiological phenotypes.

| Conductance Range | IKr | IKs | INaL | INa | ICaL |
| --- | --- | --- | --- | --- | --- |
| **Low** | 0.85 | 0.8 | 0.8 | 0.8 | 0.8 |
| **High** | 1.01 | 1.05 | 1.2 | 1.2 | 1.2 |

#### 2.4.2. Diurnal Hormonal Variations

Parting from the population described as a reference (Aguado-Sierra et al., 2024) and based on the experimentally observed effects of physiological concentrations of sex steroid hormones on ionic channels, conductances of model ion currents were modified extrapolating the methodology of Yang and Clancy (2012) to reproduce the male and female action potential duration changes after the exposure to a variety of concentration levels to testosterone, progesterone and estrogen.

A baseline population that includes a range of ages for males and various stages of the menstrual cycle for females was established. Specifically, males were categorized into three age groups (adolescence, adulthood, and senescence) and females into three hormonal stages of the menstrual cycle (early follicular phase, late follicular phase, and luteal phase). For males, application of 3 physiological concentrations of testosterone was performed to mimic the decrease of testosterone with ageing: 10, 22.5 and 35 nM reflecting normal average values measured for senescence, adulthood and adolescence (Dorgan et al., 2002). For females, the combined effects of 17*β*-estradiol (E2) and progesterone during the early follicular, late follicular, and luteal phases of the menstrual cycle were applied as: E2=0.1 nM and progesterone=2.5 nM, during the early follicular stage; E2=1.0 nM and progesterone=2.5 nM, during the late follicular stage; and E2=0.7 nM and progesterone=40.6 nM, during the luteal phase (Janse de Jonge et al., 2001). Conductance values scaling factors suggested by Yang and Clancy (Yang and Clancy, 2012) were employed for defining these baseline ages and hormonal stages in the model. Average adult testosterone conductances were extrapolated setting them to the average between senescent and adolescent reported values. This constitutes a population of 192 baseline individuals (32 subjects ∗ 3 age ranges in males + 3 menstrual hormonal stages in females), with equal representation of every hormonal stage in the population.

To apply the diurnal hormonal variation on the previously described population, 24-hour sex steroid hormones fluctuations were introduced to reproduce morning, afternoon and evening measurements. In men, there is an average daily variation of testosterone ranging from a ratio 1 (morning) to 0.5 (evening) depending on age and ethnicity, as shown by Brambilla et al (Brambilla et al., 2009) and Jain (Jain, 2015). In the case of women, estradiol and progesterone seem to vary within the day (Rahman et al., 2019; Bao et al., 2003; Patil et al., 2023), but the pattern is dependent of the menstrual cycle phase where follicular stage presents a higher fluctuation of both hormones in comparison to the luteal stage. The approximated concentrations for each of these sex hormones throughout the daytime, according to age (for men) or menstrual cycle phase (for women) are shown in Figure 2.

**Figure 2:**
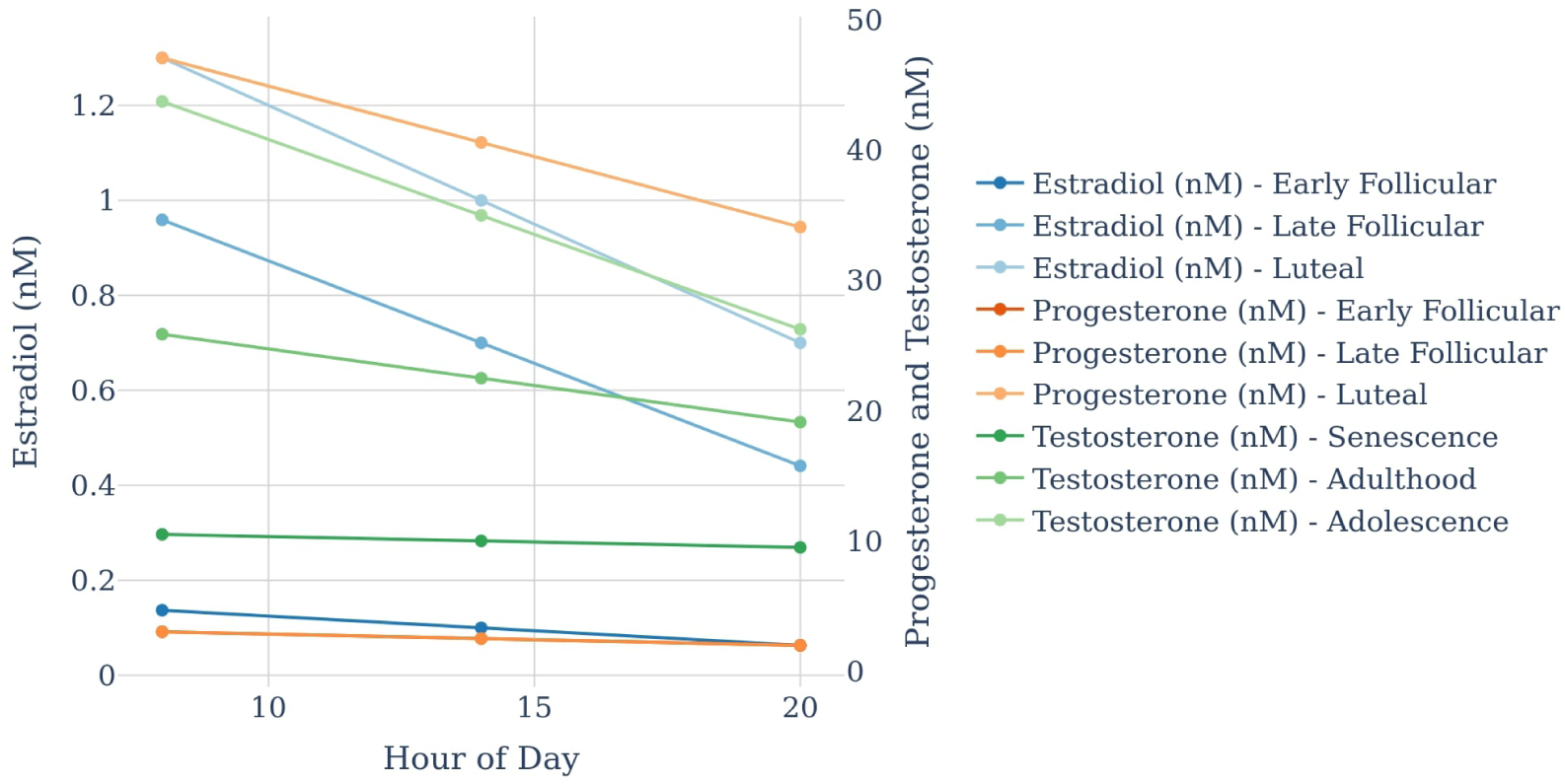
Average sex hormones concentrations (progesterone, estradiol and testosterone) decrease along daytime according to age for males and to menstrual cycle phase for females.

To introduce diurnal hormonal variations, samples were taken at three time points throughout the day (morning, afternoon, and evening) for each of the six hormonal stages (three for females and three for males) previously described. Conductance values (Tables A.7, A.8 and A.9, in Appendix A) were calculated by linearly extrapolating the experimental measurements of Yang and Clancy (2012) to provide an approximate rate of change of conductance values at the hormone concentrations of interest. Hormonal concentrations stated in Yang and Clancy were considered as afternoon average values for each hormonal stage, hence diurnal ratios were applied over them, as supported by reported hormonal concentrations with menstrual cycle for female (Janse de Jonge et al., 2001; Anckaert et al., 2021) and with age for male (Dorgan et al., 2002; Feldman et al., 2002).

As a result, 6 hormonal distinct individual stages (of males and females) were produced with sampling at 3 different daily time points: morning, afternoon and evening. Morning is considered from 7 to 12 pm, afternoon from 12 to 17 pm and evening from 17 to 22 pm.

### 2.5. Dosing Protocol

In this study the virtual population was administered moxifloxacin^2^ and dofetilide^3^, with a daily gradually escalated dose administration during 2 days (moxifloxacin: 1500 and 4500 ng/mL; dofetilide: 0.4 and 0.9 ng/mL). To mimic the plasma concentration time course after each drug dose intake, the decay in plasma concentration was estimated at each average daily time point from Figure 1 published by Darpo et al. (2015). Table 3 shows the reproduced plasma concentrations of these drugs.

**Table 3:** Drug plasma concentration at different time points applied to the computational trial.

|  |  | Moxifloxacin<br>(ng/mL) | Dofetilide<br>(ng/mL) |
| --- | --- | --- | --- |
| Dose 1 | Morning | 1500 | 0.4 |
|  | Afternoon | 900 | 0.36 |
|  | Evening | 600 | 0.2 |
| Dose 2 | Morning | 4500 | 0.9 |
|  | Afternoon | 2500 | 0.78 |
|  | Evening | 1400 | 0.38 |

### 2.6. Drug Pore Block Model

The effects of the drug affecting the ion channel conductances were incorporated based on the methodology of Mirams et al. (2011b) using a multi-channel conductance-block formulation as previously described (Aguado-Sierra et al., 2024). Table 4 shows the ion channel inhibition data used in the study.

**Table 4:** Parameters of test drugs.

| Drug |  | Cav1.2 | hERG | Kir2.1 | Kv4.3 | KvLQT1/minK | Nav1.5-late | Nav1.5-peak | Drug data |
| --- | --- | --- | --- | --- | --- | --- | --- | --- | --- |
| Moxifloxacin | IC50 | - | 93041 | - | - | 50321 | 382337 | - | Crumb et al. (2016) |
|  | h | - | 0.6 | - | - | 1.0 | 1.1 | - |  |
| Dofetilide | IC50 | - | 4.9 | - | 18.8 | - | - | - | Dutta et al. (2017) |
|  | h | - | 0.9 | - | 0.8 | - | - | - |  |

### 2.7. Pseudo-ECG Calculation

Pseudo-ECGs were calculated as detailed previously (Aguado-Sierra et al., 2024).

Briefly, the pseudo-ECG was employed to obtain markers, such as QRS and QTc. Data was analyzed using RStudio^4^ and Python^5^. An automatic algorithm was used to assess QTc interval duration, which involved identifying the segment spanning from the onset of the QRS complex to the termination of the T wave of the final simulated beat.

### 2.8. Placebo-corrected Baseline-adjusted QTc Interval

Simulations were computed for the 192 individuals of the population at baseline with morning, afternoon, and evening time points. These produced 3 baseline values per individual, that were employed to calculate a mean baseline value for each subject, which is the standard intrasubject baseline. Then, the same baseline population was simulated after the administration of the drugs. The change from baseline (ΔQTc) was computed by subtracting the standard baseline of every subject from each QTc measurement after drug intake of the corresponding subject. Placebo-corrected change from baseline (ΔΔQTc) was computed by subtracting the mean placebo ΔQTc for every timepoint from every ΔQTc measurement of the corresponding timepoint (morning, afternoon, and evening).

### 2.9. Data Analysis in Drug Risk Evaluation

Linear mixed effect (LME) Concentration-QTc (C-QTc) models are used as the analysis recommended for assessing the QTc interval prolongation risk of new drugs in early phase clinical pharmacology and tQTc studies (Garnett et al., 2018). Linear mixed models extend simple linear models by incorporating both fixed and random effects. They are particularly useful for addressing non-independence in data, which often occurs in hierarchical structures. Fixed effects are the coefficients (intercept, slope) and random effects are the variances of the intercepts or slopes across groups. These models, despite being over-parameterized, appropriately address the overall modeling objective in comparison to simple linear models. The prespecified LME C-QTc model (Eq. 1) includes ΔQTc (or ΔΔQTc) as the dependent variable, for which the fixed effect parameters are intercept, slope, influence of baseline on intercept, treatment, and nominal time from first dose. Subject identity is present as a random effect parameter on both intercept and slope terms.

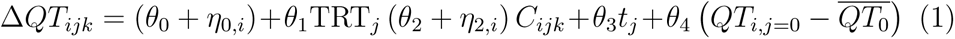

where, Δ*QT_ijk_* is the change from baseline in QTc for subject *i* in treatment *j* at time *k*; *θ*_0_ is the population mean intercept in the absence of a treatment effect; *η*_0*,i*_ is the random effect associated with the intercept term *θ*_0_; *θ*_1_ is the fixed effect associated with treatment TRT*_j_*(0= placebo, 1=active drug); *θ*_2_ is the population mean slope of the assumed linear association between concentration and Δ*QT_ijk_*; *η*_2*,i*_ is the random effect associated with the slope *θ*_2_; *C_ijk_* is the concentration for subject *i* in treatment *j* and time k; *θ*_3_ is the fixed effect associated with time; and *θ*_4_ is the fixed effect associated with baseline *QT_i,j_*_=0_; 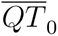 is overall mean of *QT_ij_*_0_, the mean of all the baseline QTc values at time t=0.

The term “critical concentration” denotes the concentration at which a drug induces an average increase on the QTc interval of 10 ms. These critical concentrations were determined from both clinical and computational datasets using the established definition.

### 2.10. Comparison with a Placebo Statistical Model

This investigation draws upon the analyses conducted by Minocha et al. (2019), who compiled and analyzed longitudinal data from 1,035 healthy volunteers exposed to placebo across 65 single-dose and multiple-dose Phase I trials. Employing a nonlinear mixed-effects modeling approach, Minocha et al. developed independent models for various physiological variables, including the QTc interval, to discern the influence of circadian rhythms and other covariates on placebo-induced responses.

Specifically, a dedicated nonlinear mixed-effects model was constructed for the QTc interval. The model was tailored to capture the variability in QTc interval measurements observed among the study participants following placebo administration. To account for circadian oscillations in QTc interval duration, combinations of cosine functions within the model were applied. This enabled the characterization of diurnal variations in QTc interval duration, thereby quantifying the impact of circadian rhythms on cardiac electrophysiology. Moreover, the QTc interval model incorporated significant covariates identified through the analysis, including gender, race and age. These covariates were found to contribute significantly to variability in baseline QTc interval measures, and their quantitative effects were meticulously characterized within the model.

This model provides a comprehensive framework for understanding the dynamic regulation of QTc interval duration in response to placebo and offers valuable insights into the complex interplay between circadian rhythms, demographic factors, and placebo-induced changes in cardiac electrophysiology. The concordance between the two approaches was evaluated by comparing the results generated by the in silico placebo simulations with the placebo effects predicted by the statistical model (SM).

### 2.11. Comparison with a Clinical Trial

To validate the model, the calculated placebo effect on the QTc interval (ΔΔQTc or placebo-corrected ΔQTc) was compared with results from the IQ-CSRC study by Darpo et al. (2015). The QTc interval distribution from the clinical population is shown in Figure 3. This clinical trial was performed using an incomplete block design with 9 subjects on each active drug and 6 on placebo in a parallel designed study. Subjects received a mildly prolonging dose of a well characterized drug on Day 1 and a higher dose on Day 2, or placebo. Administered doses resulted in maximum mean plasma concentrations of 1,929 ng/mL and 4,663 ng/mL for moxifloxacin and 0.43 ng/mL and 0.92 ng/mL for dofetilide, during Day 1 and Day 2 respectively. Continuous digital 12-lead ECGs were recorded at baseline and at serial time points from 1 hour prior to dosing on Day 1 until 24 hours after dosing on Day 2.

**Figure 3:**
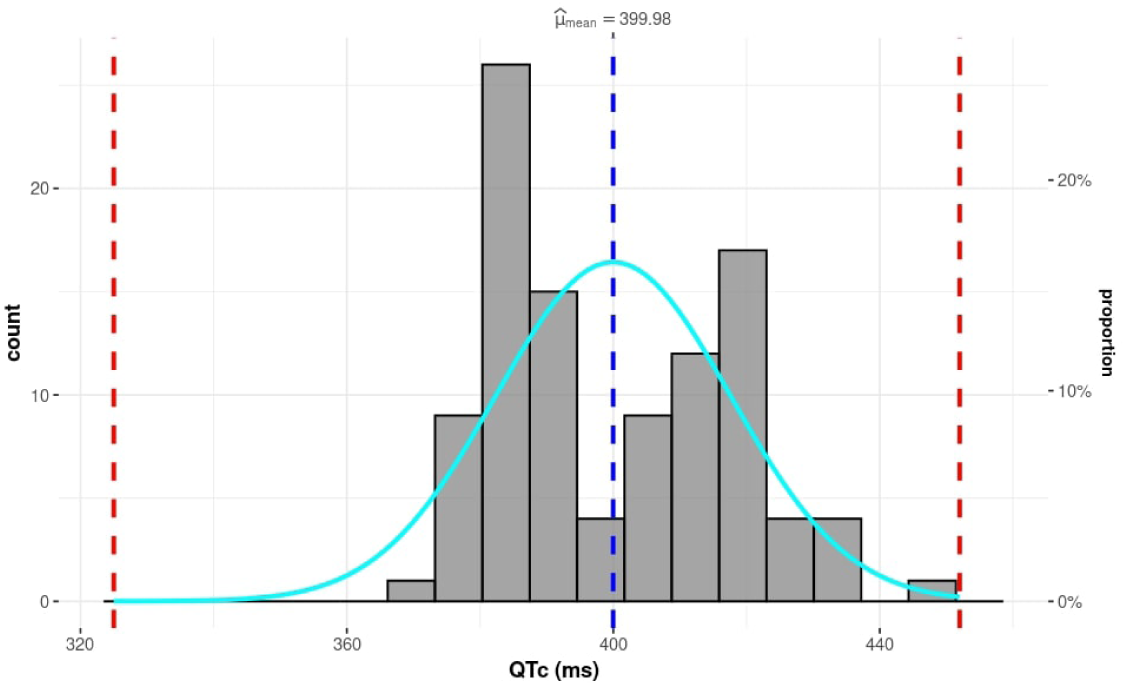
Histogram showing the distribution of the QTc intervals in the clinical population under placebo effect. Red dashed lines represent the bounds of normal ranges and the blue dashed line stands for the mean QTc value of the population.

### 2.12. Population Matching with Clinical Trials

In the moxifloxacin study group, there was one female participant among the 15 subjects, while the dofetilide group included two women out of 15. To address sex differences in our analysis, we matched the populations by selecting 15 random subjects from the virtual cohort that maintained the same sex ratio.

### 2.13. Computational Requirements

This work required an approximate 14 million core-hours of computation in the Vega Supercomputer ^6^, a Slovenian petascale supercomputer with a sustained performance of 6.9 petaflops. A total number of 2,880 full heart simulations were performed under the scope of this project for the final data analysis.

## 3. Results

### 3.1. Characterization of QTc Interval in the Virtual Population

The QTc placebo distribution of the population of 192 individuals is shown in Figure 4. Notably, it falls within normal ranges in humans (Mason et al., 2007). However, there were a few female QTc interval values slightly outside the normal bounds, but still in the clinically accepted borderline range in women. These subjects were not excluded from the study.

**Figure 4:**
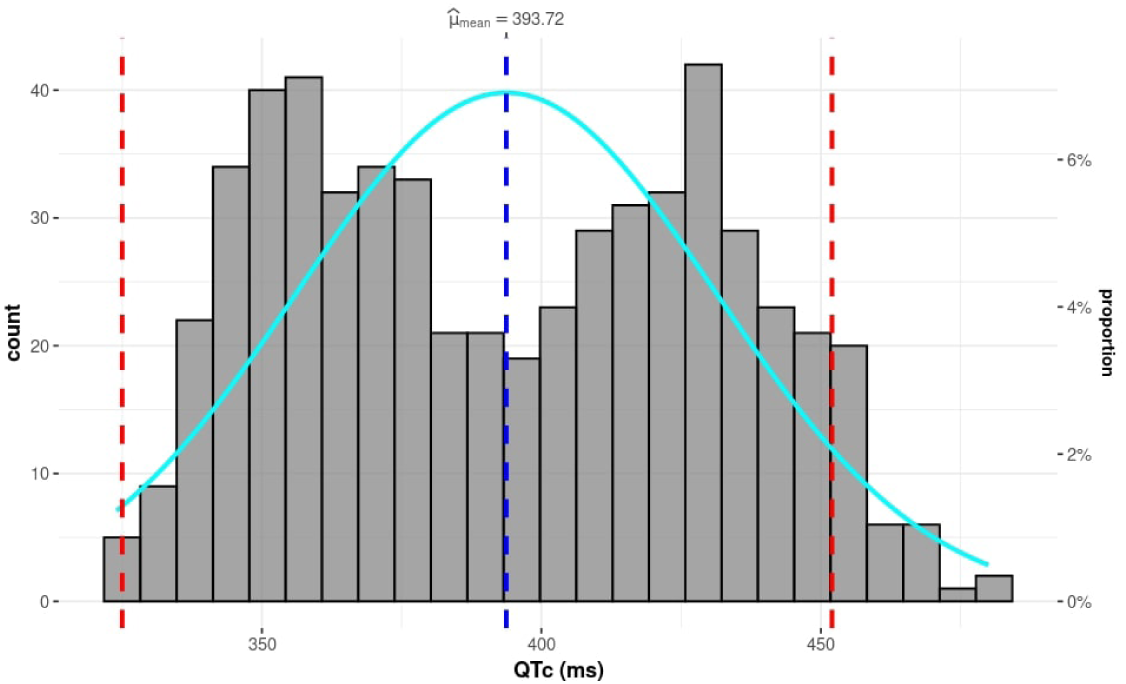
Histograms showing the distribution of the QTc interval of the in silico placebo population. Red dashed lines represent the bounds of normal ranges and the blue dashed line indicates the mean QTc value of the population.

### 3.2. Validation of the Computational Placebo Effect

The SM was used to compute the variability of QTc interval measurements for every subject. ΔQTc measurements were computed by quantifying the difference between the simulated QTc intervals and the estimated QTc oscillations derived from the SM for each individual participant. This enabled a comparison of the variability observed in the computational data, the SM-enhanced computational data, and the clinical data.

The results of this analysis, shown in Figure 5, revealed similar distributions of ΔQTc measurements between the three trial datasets. Moreover, when comparing the computational trial with and without the SM, it is observed that the computational trial with the diurnal hormonal variations was able to capture approximately 50% of the variability observed in clinical trials. This suggests that this approach, which considers hormonal changes and circadian rhythms, effectively captures one of the the main factors associated with the spectrum of placebo-induced QTc interval variability reported in clinical settings.

**Figure 5:**
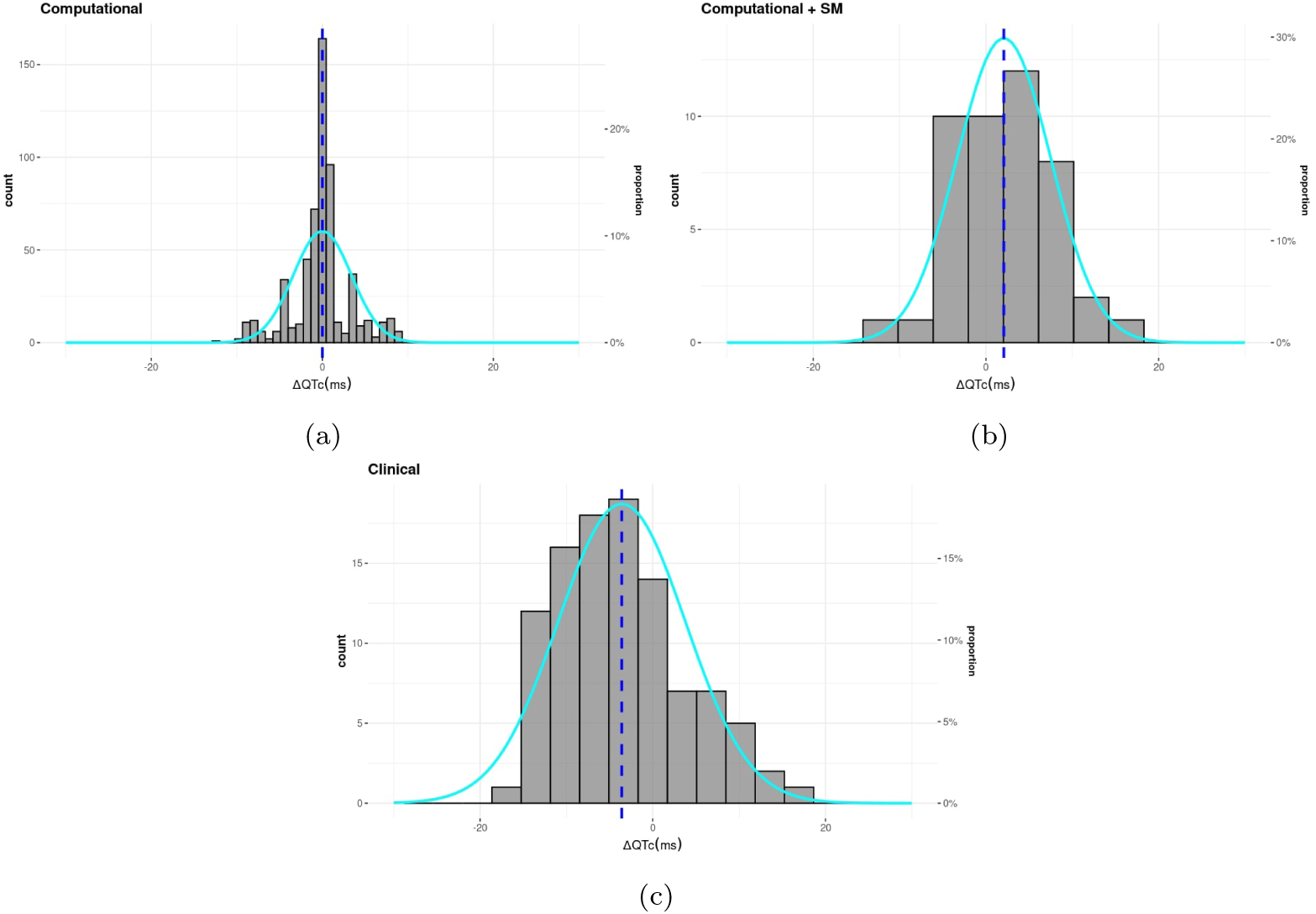
Histograms showing the distribution of the placebo ΔQTc in the computational (a), SM-enhanced computational (b) and clinical trials (c).

To further quantify the differences between these distributions, the Total Variation Distance (TVD) between the ΔQTc distributions in the clinical trial, the computational trial, and SM-enhanced computational trial were calculated. The TVD between the clinical trial and computational trial was 0.4439, indicating a moderate to high level of dissimilarity, suggesting notable differences in ΔQTc values. In contrast, the TVD between the clinical trial and the SM-enhanced computational trial was much lower, at 0.0848, reflecting a strong similarity and substantial overlap between these distributions. The TVD between the computational trial and the SM-enhanced computational trial was 0.4405, further underscoring the difference introduced by the SM. These findings highlight the effectiveness of the SM in capturing ΔQTc variability, aligning the computational trial more closely with the observed clinical data and providing confidence in the methodology, while also suggesting room for the inclusion of other placebo effects currently not taken into account in the computational trial.

### 3.3. Comparing the Computational and the Clinical Trial

Figures 6 and 7 show the C-QTc model and the ΔΔQTc and plasma concentration by timepoint for both the computational and clinical trials for moxifloxacin and dofetilide, respectively.

**Figure 6:**
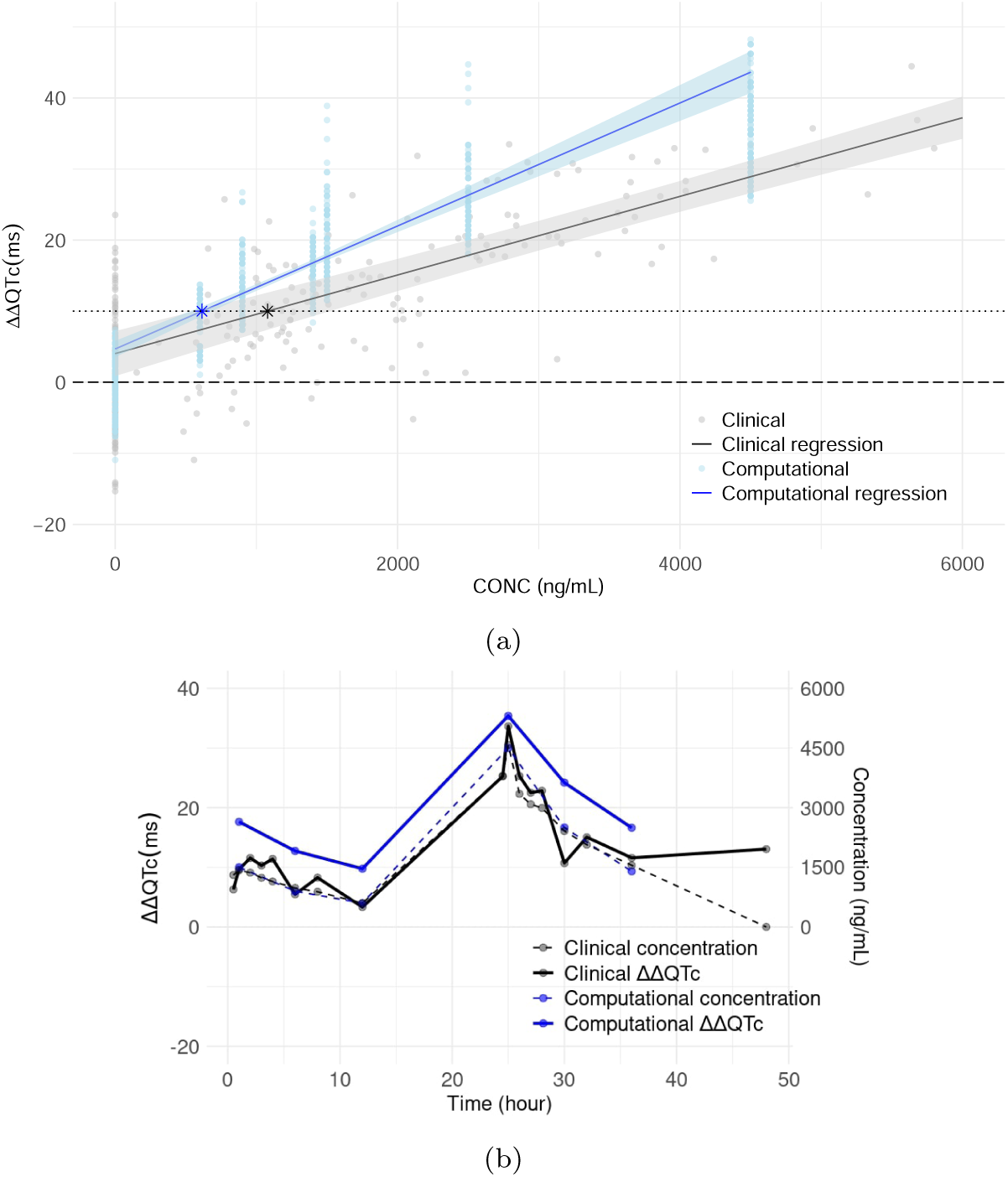
(a) Predicted effect on ΔΔQTc using C-QTc effect models in the clinical and computational trials for moxifloxacin. The regression lines include stars to indicate their predicted critical concentration (at the 10 ms threshold). (b) Observed ΔΔQTc (solid lines, left y-axis) and plasma concentration (dashed lines, right y-axis) by timepoint on day 1 and day 2 in the clinical and computational trials for moxifloxacin.

**Figure 7:**
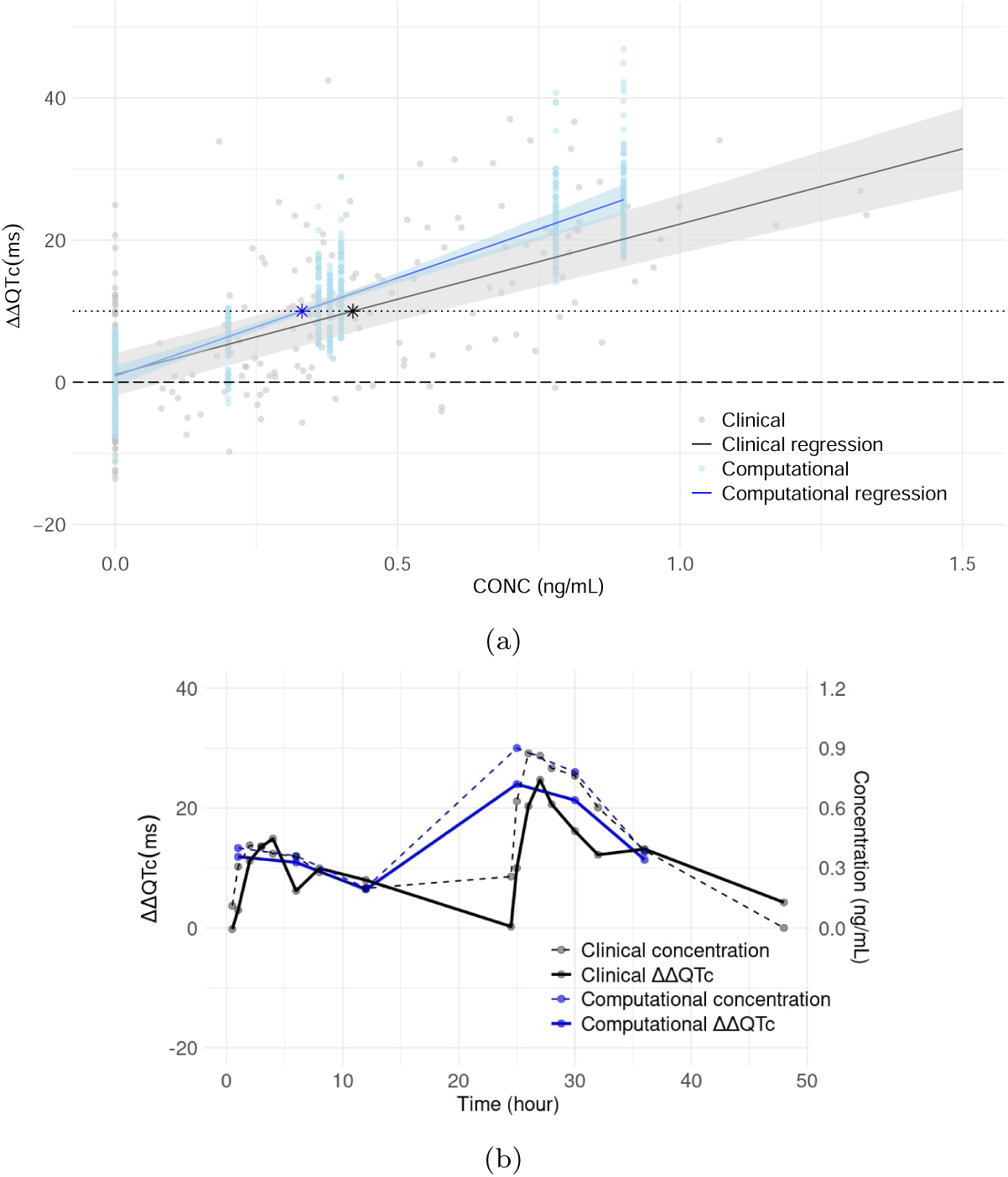
(a) Predicted effect on ΔΔQTc using C-QTc effect models in the clinical and computational trials for dofetilide. The regression lines include stars to indicate their predicted critical concentration (at the 10 ms threshold). (b) Observed ΔΔQTc (solid lines, left y-axis) and plasma concentration (dashed lines, right y-axis) by timepoint on day 1 and day 2 in the clinical and computational trials for dofetilide.

The critical concentrations observed on the C-QTc effect models of Figures 6(a) and 7(a) are reported along with the most relevant statistical data in Tables 5 and 6. The computational C-QTc model resulted in smaller critical concentration values both for moxifloxacin and dofetilide in comparison to the clinical model, which had already been observed by Aguado-Sierra et al. (2024). Moxifloxacin, in particular, showed the greatest discrepancy, with concentrations being 2.5 times lower than those reported in clinical studies. Additionally, the slopes demonstrated relative errors ranging from 30% to 56%, consistent with observed interstudy variability (Lin et al., 2021), and intersectant confidence intervals.

**Table 5:** Slope and intercept of concentration-QTc model and associated metrics estimated from the computational and clinical data of Figures 6(a) and 7(a).

|  |  |  | Coef | Std err | t | 95.0% Conf. Interval | Relative error of slopes | Relative error of intercepts |
| --- | --- | --- | --- | --- | --- | --- | --- | --- |
| Moxifloxacin | Computational | Intercept | 4.68 | 0.69 | 6.83 | [3.34, 6.03] | 56% | 61% |
|  |  | Slope | 0.0087 | 0.00054 | 16.14 | [0.0076, 0.0097] |  |  |
|  | Clinical | Intercept | 2.90 | 1.94 | 1.50 | [-0.89, 6.70] |  |  |
|  |  | Slope | 0.0055 | 0.00045 | 12.24 | [0.0046, 0.0064] |  |  |
| Dofetilide | Computational | Intercept | 0.89 | 0.85 | 1.05 | [-0.77, 2.55] | 30% | 21% |
|  |  | Slope | 27.55 | 2.31 | 11.95 | [23.03, 32.07] |  |  |
|  | Clinical | Intercept | 1.12 | 1.82 | 0.61 | [-2.46, 4.70] |  |  |
|  |  | Slope | 21.16 | 2.42 | 8.73 | [16.41, 25.91] |  |  |

Figures 6(b) and 7(b) present a comparison between computational predictions and clinical measurements for moxifloxacin and dofetilide, focusing on ΔΔQTc and drug concentration over time. For moxifloxacin, the average error in ΔΔQTc was 6.99 ms, and the mean concentration error was 78.41 ng/mL, indicating a good overall fit with clinical data. In addition, dofetilide demonstrated even greater precision, with an average ΔΔQTc error of 6.02 ms and a remarkably low concentration error of 0.065 ng/mL.

### 3.4. The Role of Sex Hormones on ΔΔQTc

The ΔQTc values obtained for every hormonal stage under placebo effect in the computational population are shown in Figure A.8. Additionally, Figures A.9 and A.10 display the ΔΔQTc values for each hormonal stage under drug effect. Sex-related differences in ΔΔQTc expression prove that the variability introduced by hormonal changes is not negligible, and foremost, how determined stages tend to present higher variability and mean ΔΔQTc effect. Table 6 shows the difference in the critical concentrations associated to each hormonal stage, which demonstrates the impact of sex steroid hormones on the phenotypic expression of QTc interval.

**Table 6:** Critical concentrations estimated from the computational and clinical data of Figures 6(a) and 7(a), also specified by hormonal stage for the computational trial.

|  |  | Critical Concentration (ng/mL) | Hormonal Stage |
| --- | --- | --- | --- |
| Moxifloxacin | Clinical | 1574 | - |
|  | Computational | 614 | - |
|  |  | 540 | Early Follicular |
|  |  | 529 | Late Follicular |
|  |  | 572 | Luteal |
|  |  | 696 | Adolescence |
|  |  | 673 | Adulthood |
|  |  | 728 | Senescence |
| Dofetilide | Clinical | 0.55 | - |
|  | Computational | 0.33 | - |
|  |  | 0.31 | Early Follicular |
|  |  | 0.30 | Late Follicular |
|  |  | 0.32 | Luteal |
|  |  | 0.35 | Adolescence |
|  |  | 0.36 | Adulthood |
|  |  | 0.35 | Senescence |

### 3.5. The role of Circadian Rhythm on ΔΔQTc

The ΔQTc values obtained at various time points throughout the day (morning, afternoon, and evening) under the placebo effect in the computational population are shown in Figure A.11. Figures A.12 and A.13 illustrate the behavior of ΔΔQTc measurements for moxifloxacin and dofetilide at various time points throughout the day in both the computational and clinical studies, demonstrating a good correlation between them.

### 3.6. Population Matching between Clinical and Computational Trials

Figure A.14 show the results from the population matching for comparison and validation of the results against the clinical trial data for both moxifloxacin and dofetilide, where both provide improved results, highlighting the bias introduced by the sex ratio of the cohort.

## 4. Discussion

This study presents a novel computational approach to assess concentration QTc-relationships using a virtual patient population with 3D heart simulations able to reproduce a full human clinical trial paradigm. The methodology to reproduce a normal population incorporated a realistic sex-specific anatomical description, sex-specific phenotypes based on differences in ion channel conductances, and approximately accounted for normal intersubject and intrasubject electrophysiological phenotype variability introduced by sex steroid hormonal changes. These also included diurnal mechanistic effects to assess the extent of their contribution to the placebo effect.

To replicate clinical trial results using a LME model, the computational trial must include a sufficient number of samples to ensure model convergence. Accounting for diurnal hormonal variation stages addresses the sample size requirement and enables the definition of appropriate timepoints for drug intake, as advised by regulatory bodies. Furthermore, incorporating various hormonal stages enhances the accuracy of reproducing placebo-related ΔQT variability, which is essential for matching the C-QTc model used in clinical trials.

The QTc-interval values (Figure 4) and QTc-interval prolongations (Figure 5) obtained from the computational framework fell within the confidence intervals reported in Mason et al. (2007) and Darpo et al. (2015) studies, respectively. Results show that employing placebo-corrected ΔQTc in computational trials, produced by circadian rhythm variations simplified as diurnal hormonal changes, provides a good correlation with the large population-based SM and the clinical trial data. Despite showing a more reduced range of variability in comparison to clinical measurements, this work demonstrates that hormonal changes play an important role in circadian rhythm. Nevertheless, the hormonal changes performed were conservative, and a larger range of hormonal variations could increase the observed diurnal effect.

The sex-related and circadian rhythm influence was well approximated as compared to both, the expected behavior from experimental studies and the clinical trial data as observed in Figures A.9, A.10, A.12 and A.13. Sex variations show the prevalence of larger variability and longer ΔΔQTc values in women in comparison to men. Moreover, higher ΔΔQTc values were observed for women in the luteal and late follicular phases. Results regarding the increasing ΔΔQTc values with ageing in men (according to the decrease in serum testosterone) were inconclusive, despite the slightly higher C-QTc slopes reported for the adolescent and senescent cohorts. Daily variations present wider ranges in the morning, with a decreasing tendency of ΔΔQTc values along the day in agreement with the decrease in plasma concentration after drug intake, both in the computational and clinical trial.

There are many sources of variability in the drug characterization process that represent a hurdle for the assessment of drug-induced arrhythmia both clinically and computationally. This variability is leading to challenges in defining adequate dose regimens in clinical trials for drug safety. One example is variability related to experimental high-throughput ion channel electrophysiology screenings. Uncertainty in the Hill coefficient (h) and the half-maximal inhibitory concentration (IC50) of reported concentration-effect curves is particularly high. Furthermore, depending on a compound’s ion channel blocking profile, the uncertainty introduced may become relevant (Elkins et al., 2013). A similar problem happens with the fraction unbound in plasma, which is an important parameter with high intersubject variability (Wang et al., 2014). In addition, experimental subjects taking part in clinical trials are heterogeneous beyond their electrophysiological phenotype, meaning that the variability of these populations adds “noise” in clinical trial data; which is generally a reason for the unequal distribution of female subjects in cardiac safety trials. Lastly, with respect to the placebo effect, there are several physiologic factors that may be involved to a higher or lower extent.

Interestingly, the computational matched population results exhibited an increased predictive accuracy including both, the computational placebo and the statistical placebo model. The latter demonstrated a notable improvement in comparison to the clinical trial for both compounds (Figure A.14). This significant improvement highlights the importance of addressing clinical biases and demographic factors in drug safety studies. Furthermore, it underscores the added value of the placebo effect, providing stronger credibility to the computational approach.

The use of in silico human clinical trials for cardiac safety drug testing is of great interest because they allow fast and affordable assessment of any plasma concentration of a given drug on cardiac electrophysiology and provides results directly comparable to clinical trials. This may be particularly valuable for oncology drugs with poor tolerability that limits doses and exposures above the intended clinical dose. The full replication of a clinical trial, including the placebo effect, provides further confidence on the model to reproduce normal human physiological behavior during baseline and after the administration of potentially QTc prolonging drugs.

An important question remains: Is an in silico placebo population necessary to reproduce a human clinical trial? Results show little difference between full in silico clinical trial (Figures 6 and 7) when comparing placebo-corrected ΔQTc (ΔΔQTc) with ΔQTc, as previously done (Aguado-Sierra et al., 2024). This suggests that the addition of an in silico placebo population does not significantly enhance the accuracy or alignment with clinical data. However, this approach provides the ability to apply a linear mixed-effects model, which is common practice in clinical settings.

The most interesting benefit of the new approach lies in its potential to increase the variability reproduced in the simulations, allowing for a more comprehensive study that can better capture the inherent uncertainty present in human physiology. While circadian rhythm might be closely linked to cardiovascular events, potentially allowing the models and assumptions presented here to shed light on their origins, this expanded variability enables a deeper exploration of the nuances in human responses, even though no substantial improvements or discrepancies were observed when comparing the current findings with those from previous work.

### 4.1. Conclusions

This framework can be employed to assess the concentration-ΔΔQTc relationship of a given drug on a virtual human population, which directly translates into human clinical trial data. It does also reduce the limitations of 0-Dimensional (0D) models, that are not able to reproduce clinically observed markers related to organ-level interactions such as QTc-interval prolongation. This opens the opportunity to perform QTc assessment based on concentration-ΔΔQTc in silico modeling in the drug discovery phase, thereby allowing a much earlier understanding of potential QTc effects as compared to clinical data. Computational trials offer an unlimited dosage testing range, widening the opportunities to assess more accurately the optimal dosage and adapting to the specific trial needs without the risk and cost-related burden of current procedures.

This approach is intended to provide reliable predictions of the risk of developing arrhythmia in a realistic sex-specific heart population which reflects human placebo-induced variability, and therefore, provides predictions closer to clinical trial outcomes. This study provides evidence of the role of hormone variations in the placebo effect observed in tQTc studies, which is key to accurately reproduce the gold standard biomarkers in proarrhythmic assessment and forecast drug evolution in alignment with regulatory agencies’ guidelines.

Moreover, this work opens a new paradigm in addressing clinical biases by highlighting the relevance of human diversity and the need to include underrepresented groups in these studies. Including sex-specific differences underscored the importance of variability in clinical studies, ensuring more accurate and representative assessments.

### 4.2. Limitations and Future Work

As discussed previously, the fraction unbound and ion channel IC50 data are two relevant parameters that have a high impact on the results of the model. To address this uncertainty, providing confidence intervals for these parameters could offer a better understanding of their effects within the proposed models. This is one of the main hurdles for in silico trials, as there are limitations of quality preclinical data.

Another limitation to the electrophysiology simulations in this study is the lack of electromechanical simulations that could provide mechanistic information regarding the hemodynamic effects of these drugs. An extension to the solution of an electromechanical model to assess drugs in a large population would require approximately 6.5 times more computation time. While running electromechanics was not an aim of this work, it constitutes a part of future work.

It is important to highlight the high computational demand associated to this type of study. Nevertheless, computational time demand on commercial and research HPC infrastructures is increasing constantly and it will not be a limiting factor to in silico human heart trials. Therefore, provided the adequate platform and computational time accessibility, these workflows could be integrated within the drug development pipeline.

Finally, further key placebo-related factors could be included in the virtual population to broaden the electrocardiographic phenotype expression, and therefore get a step closer to mechanistically reproduce the clinical spectrum. Future studies could leverage smartwatch data, which offers continuous day-long monitoring. This would enable a comprehensive analysis of the relationship between any recorded biometric data and electrocardiographic responses.

## Data Availability

The methodology can be replicated using any finite element solver given all the parameterization information provided in this paper. Quantified biomarkers can be made available upon request.

## Acknowledgements

This work was supported by the Partnership for Advanced Computing in Europe (grant number EHPC-REG-2022R01-038-EuroHPC) awarded to J.A.

## Authorship Contribution Statement

**Paula Dominguez-Gomez**: Conceptualization, Data curation, Formal analysis, Investigation, Methodology, Project administration, Resources, Validation, Visualization, Writing - original draft, Writing - review & editing. **Constantine Butakoff** : Conceptualization, Data curation, Formal analysis, Investigation, Methodology, Software, Supervision, Validation, Visualization, Writing - original draft, Writing - review & editing. **Georg Rast**: Conceptualization, Data curation, Supervision, Validation, Writing - original draft, Writing - review & editing. **Borje Darpo**: Conceptualization, Data curation, Resources, Supervision, Validation, Visualization, Writing - original draft, Writing - review & editing. **Mariano Vazquez**: Funding acquisition, Project administration, Resources, Software, Supervision, Writing - review & editing. **Jazmin Aguado-Sierra**: Conceptualization, Data curation, Formal analysis, Funding acquisition, Investigation, Methodology, Project administration, Resources, Software, Supervision, Validation, Visualization, Writing - original draft, Writing - review & editing.

## Declarations of Competing Insterest

M.V. is CTO and co-founder of ELEM Biotech. Elem Biotech owns the commercial rights to Alya, the computational finite element solver employed in this study.

## Appendix A. Supplementary Material

**Table A.7:**
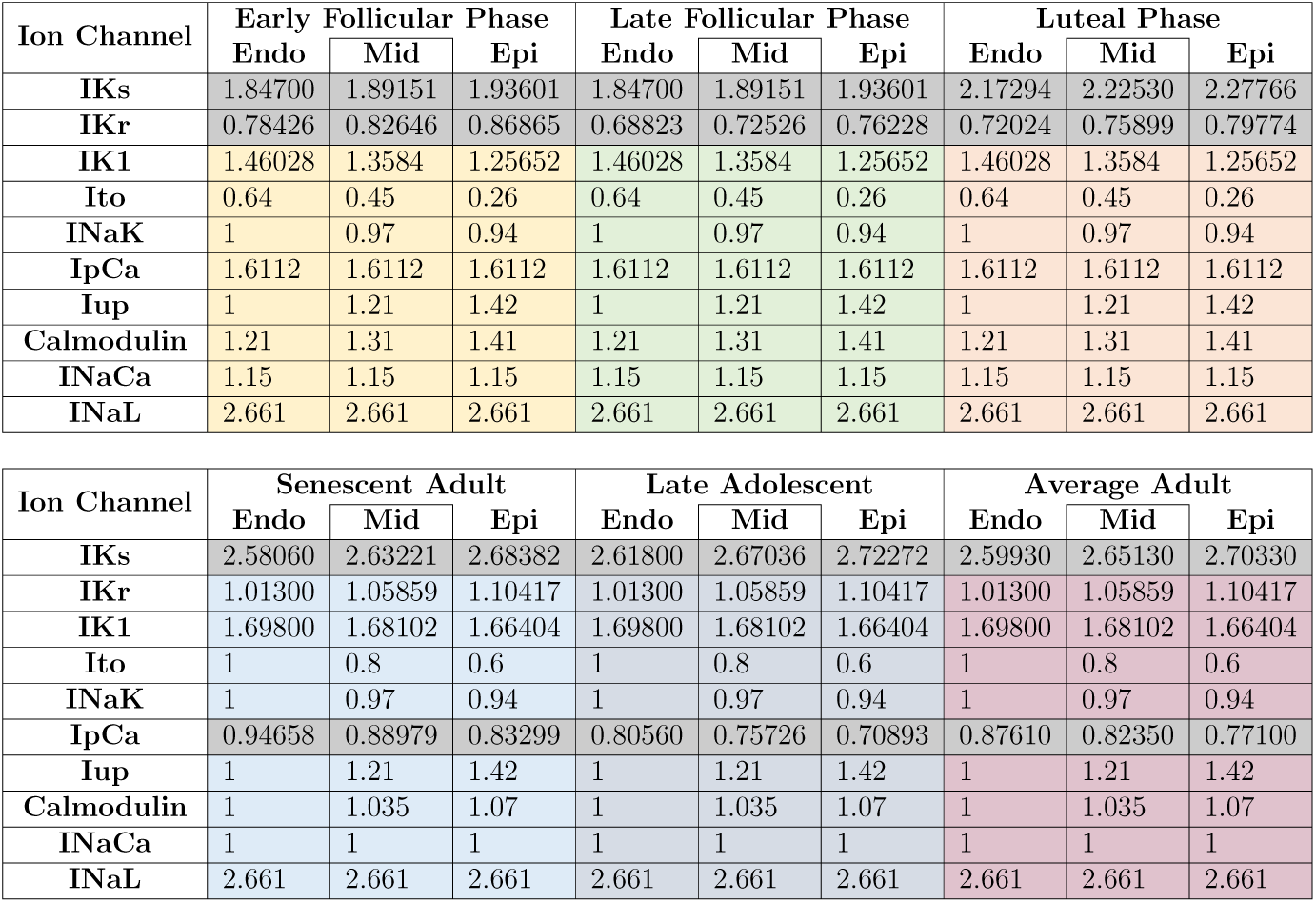
Sex-specific baseline conductance differences for afternoon hormonal changes. Grey shaded areas stand for channels affected by each hormonal change.

| Ion Channel | Early Follicular Phase |  |  | Late Follicular Phase |  |  | Luteal Phase |  |  |
| --- | --- | --- | --- | --- | --- | --- | --- | --- | --- |
|  | Endo | Mid | Epi | Endo | Mid | Epi | Endo | Mid | Epi |
| IKs | 1.84700 | 1.89151 | 1.93601 | 1.84700 | 1.89151 | 1.93601 | 2.17294 | 2.22530 | 2.27766 |
| IKr | 0.78426 | 0.82646 | 0.86865 | 0.68823 | 0.72526 | 0.76228 | 0.72024 | 0.75899 | 0.79774 |
| IK1 | 1.46028 | 1.3584 | 1.25652 | 1.46028 | 1.3584 | 1.25652 | 1.46028 | 1.3584 | 1.25652 |
| Ito | 0.64 | 0.45 | 0.26 | 0.64 | 0.45 | 0.26 | 0.64 | 0.45 | 0.26 |
| INaK | 1 | 0.97 | 0.94 | 1 | 0.97 | 0.94 | 1 | 0.97 | 0.94 |
| IpCa | 1.6112 | 1.6112 | 1.6112 | 1.6112 | 1.6112 | 1.6112 | 1.6112 | 1.6112 | 1.6112 |
| Iup | 1 | 1.21 | 1.42 | 1 | 1.21 | 1.42 | 1 | 1.21 | 1.42 |
| Calmodulin | 1.21 | 1.31 | 1.41 | 1.21 | 1.31 | 1.41 | 1.21 | 1.31 | 1.41 |
| INaCa | 1.15 | 1.15 | 1.15 | 1.15 | 1.15 | 1.15 | 1.15 | 1.15 | 1.15 |
| INaL | 2.661 | 2.661 | 2.661 | 2.661 | 2.661 | 2.661 | 2.661 | 2.661 | 2.661 |

| Ion Channel | Senescent Adult |  |  | Late Adolescent |  |  | Average Adult |  |  |
| --- | --- | --- | --- | --- | --- | --- | --- | --- | --- |
|  | Endo | Mid | Epi | Endo | Mid | Epi | Endo | Mid | Epi |
| IKs | 2.58060 | 2.63221 | 2.68382 | 2.61800 | 2.67036 | 2.72272 | 2.59930 | 2.65130 | 2.70330 |
| IKr | 1.01300 | 1.05859 | 1.10417 | 1.01300 | 1.05859 | 1.10417 | 1.01300 | 1.05859 | 1.10417 |
| IK1 | 1.69800 | 1.68102 | 1.66404 | 1.69800 | 1.68102 | 1.66404 | 1.69800 | 1.68102 | 1.66404 |
| Ito | 1 | 0.8 | 0.6 | 1 | 0.8 | 0.6 | 1 | 0.8 | 0.6 |
| INaK | 1 | 0.97 | 0.94 | 1 | 0.97 | 0.94 | 1 | 0.97 | 0.94 |
| IpCa | 0.94658 | 0.88979 | 0.83299 | 0.80560 | 0.75726 | 0.70893 | 0.87610 | 0.82350 | 0.77100 |
| Iup | 1 | 1.21 | 1.42 | 1 | 1.21 | 1.42 | 1 | 1.21 | 1.42 |
| Calmodulin | 1 | 1.035 | 1.07 | 1 | 1.035 | 1.07 | 1 | 1.035 | 1.07 |
| INaCa | 1 | 1 | 1 | 1 | 1 | 1 | 1 | 1 | 1 |
| INaL | 2.661 | 2.661 | 2.661 | 2.661 | 2.661 | 2.661 | 2.661 | 2.661 | 2.661 |

**Table A.8:** Sex-specific baseline conductance differences for morning hormonal changes. Darker shaded areas stand for channels affected by morning hormonal changes.

| Ion Channel | Early Follicular Phase |  |  | Late Follicular Phase |  |  | Luteal Phase |  |  |
| --- | --- | --- | --- | --- | --- | --- | --- | --- | --- |
|  | Endo | Mid | Epi | Endo | Mid | Epi | Endo | Mid | Epi |
| IKs | 1.85148 | 1.89610 | 1.94071 | 1.85148 | 1.89610 | 1.94071 | 2.22838 | 2.28208 | 2.33577 |
| IKr | 0.78029 | 0.82227 | 0.86425 | 0.69261 | 0.72987 | 0.76713 | 0.65623 | 0.69153 | 0.72684 |
| IK1 | 1.46028 | 1.3584 | 1.25652 | 1.46028 | 1.3584 | 1.25652 | 1.46028 | 1.3584 | 1.25652 |
| Ito | 0.64 | 0.45 | 0.26 | 0.64 | 0.45 | 0.26 | 0.64 | 0.45 | 0.26 |
| INaK | 1 | 0.97 | 0.94 | 1 | 0.97 | 0.94 | 1 | 0.97 | 0.94 |
| IpCa | 1.6112 | 1.6112 | 1.6112 | 1.6112 | 1.6112 | 1.6112 | 1.6112 | 1.6112 | 1.6112 |
| Iup | 1 | 1.21 | 1.42 | 1 | 1.21 | 1.42 | 1 | 1.21 | 1.42 |
| Calmodulin | 1.21 | 1.31 | 1.41 | 1.21 | 1.31 | 1.41 | 1.21 | 1.31 | 1.41 |
| INaCa | 1.15 | 1.15 | 1.15 | 1.15 | 1.15 | 1.15 | 1.15 | 1.15 | 1.15 |
| INaL | 2.661 | 2.661 | 2.661 | 2.661 | 2.661 | 2.661 | 2.661 | 2.661 | 2.661 |

| Ion Channel | Senescent Adult |  |  | Late Adolescent |  |  | Average Adult |  |  |
| --- | --- | --- | --- | --- | --- | --- | --- | --- | --- |
|  | Endo | Mid | Epi | Endo | Mid | Epi | Endo | Mid | Epi |
| IKs | 2.58135 | 2.63297 | 2.68460 | 2.63109 | 2.68371 | 2.73633 | 2.60435 | 2.65644 | 2.70852 |
| IKr | 1.01300 | 1.05859 | 1.10417 | 1.01300 | 1.05859 | 1.10417 | 1.01300 | 1.05859 | 1.10417 |
| IK1 | 1.69800 | 1.68102 | 1.66404 | 1.69800 | 1.68102 | 1.66404 | 1.69800 | 1.68102 | 1.66404 |
| Ito | 1 | 0.8 | 0.6 | 1 | 0.8 | 0.6 | 1 | 0.8 | 0.6 |
| INaK | 1 | 0.97 | 0.94 | 1 | 0.97 | 0.94 | 1 | 0.97 | 0.94 |
| IpCa | 0.94376 | 0.88713 | 0.83051 | 0.75626 | 0.71088 | 0.66551 | 0.85706 | 0.80563 | 0.75421 |
| Iup | 1 | 1.21 | 1.42 | 1 | 1.21 | 1.42 | 1 | 1.21 | 1.42 |
| Calmodulin | 1 | 1.035 | 1.07 | 1 | 1.035 | 1.07 | 1 | 1.035 | 1.07 |
| INaCa | 1 | 1 | 1 | 1 | 1 | 1 | 1 | 1 | 1 |
| INaL | 2.661 | 2.661 | 2.661 | 2.661 | 2.661 | 2.661 | 2.661 | 2.661 | 2.661 |

**Table A.9:** Sex-specific baseline conductance differences for evening hormonal changes. Darker shaded areas stand for channels affected by evening hormonal changes.

| Ion Channel | Early Follicular Phase |  |  | Late Follicular Phase |  |  | Luteal Phase |  |  |
| --- | --- | --- | --- | --- | --- | --- | --- | --- | --- |
|  | Endo | Mid | Epi | Endo | Mid | Epi | Endo | Mid | Epi |
| IKs | 1.84250 | 1.88690 | 1.93130 | 1.84250 | 1.88690 | 1.93130 | 2.11727 | 2.16829 | 2.21931 |
| IKr | 0.78819 | 0.83059 | 0.87299 | 0.74786 | 0.78810 | 0.82833 | 0.72023 | 0.75898 | 0.79773 |
| IK1 | 1.46028 | 1.3584 | 1.25652 | 1.46028 | 1.3584 | 1.25652 | 1.46028 | 1.3584 | 1.25652 |
| Ito | 0.64 | 0.45 | 0.26 | 0.64 | 0.45 | 0.26 | 0.64 | 0.45 | 0.26 |
| INaK | 1 | 0.97 | 0.94 | 1 | 0.97 | 0.94 | 1 | 0.97 | 0.94 |
| IpCa | 1.6112 | 1.6112 | 1.6112 | 1.6112 | 1.6112 | 1.6112 | 1.6112 | 1.6112 | 1.6112 |
| Iup | 1 | 1.21 | 1.42 | 1 | 1.21 | 1.42 | 1 | 1.21 | 1.42 |
| Calmodulin | 1.21 | 1.31 | 1.41 | 1.21 | 1.31 | 1.41 | 1.21 | 1.31 | 1.41 |
| INaCa | 1.15 | 1.15 | 1.15 | 1.15 | 1.15 | 1.15 | 1.15 | 1.15 | 1.15 |
| INaL | 2.661 | 2.661 | 2.661 | 2.661 | 2.661 | 2.661 | 2.661 | 2.661 | 2.661 |

| Ion Channel | Senescent Adult |  |  | Late Adolescent |  |  | Average Adult |  |  |
| --- | --- | --- | --- | --- | --- | --- | --- | --- | --- |
|  | Endo | Mid | Epi | Endo | Mid | Epi | Endo | Mid | Epi |
| IKs | 2.57985 | 2.63145 | 2.68305 | 2.60491 | 2.65701 | 2.70911 | 2.59425 | 2.64614 | 2.69802 |
| IKr | 1.01300 | 1.05859 | 1.10417 | 1.01300 | 1.05859 | 1.10417 | 1.01300 | 1.05859 | 1.10417 |
| IK1 | 1.69800 | 1.68102 | 1.66404 | 1.69800 | 1.68102 | 1.66404 | 1.69800 | 1.68102 | 1.66404 |
| Ito | 1 | 0.8 | 0.6 | 1 | 0.8 | 0.6 | 1 | 0.8 | 0.6 |
| INaK | 1 | 0.97 | 0.94 | 1 | 0.97 | 0.94 | 1 | 0.97 | 0.94 |
| IpCa | 0.94940 | 0.89244 | 0.83547 | 0.85494 | 0.80365 | 0.75235 | 0.89512 | 0.84141 | 0.78771 |
| Iup | 1 | 1.21 | 1.42 | 1 | 1.21 | 1.42 | 1 | 1.21 | 1.42 |
| Calmodulin | 1 | 1.035 | 1.07 | 1 | 1.035 | 1.07 | 1 | 1.035 | 1.07 |
| INaCa | 1 | 1 | 1 | 1 | 1 | 1 | 1 | 1 | 1 |
| INaL | 2.661 | 2.661 | 2.661 | 2.661 | 2.661 | 2.661 | 2.661 | 2.661 | 2.661 |

**Figure A.8:**
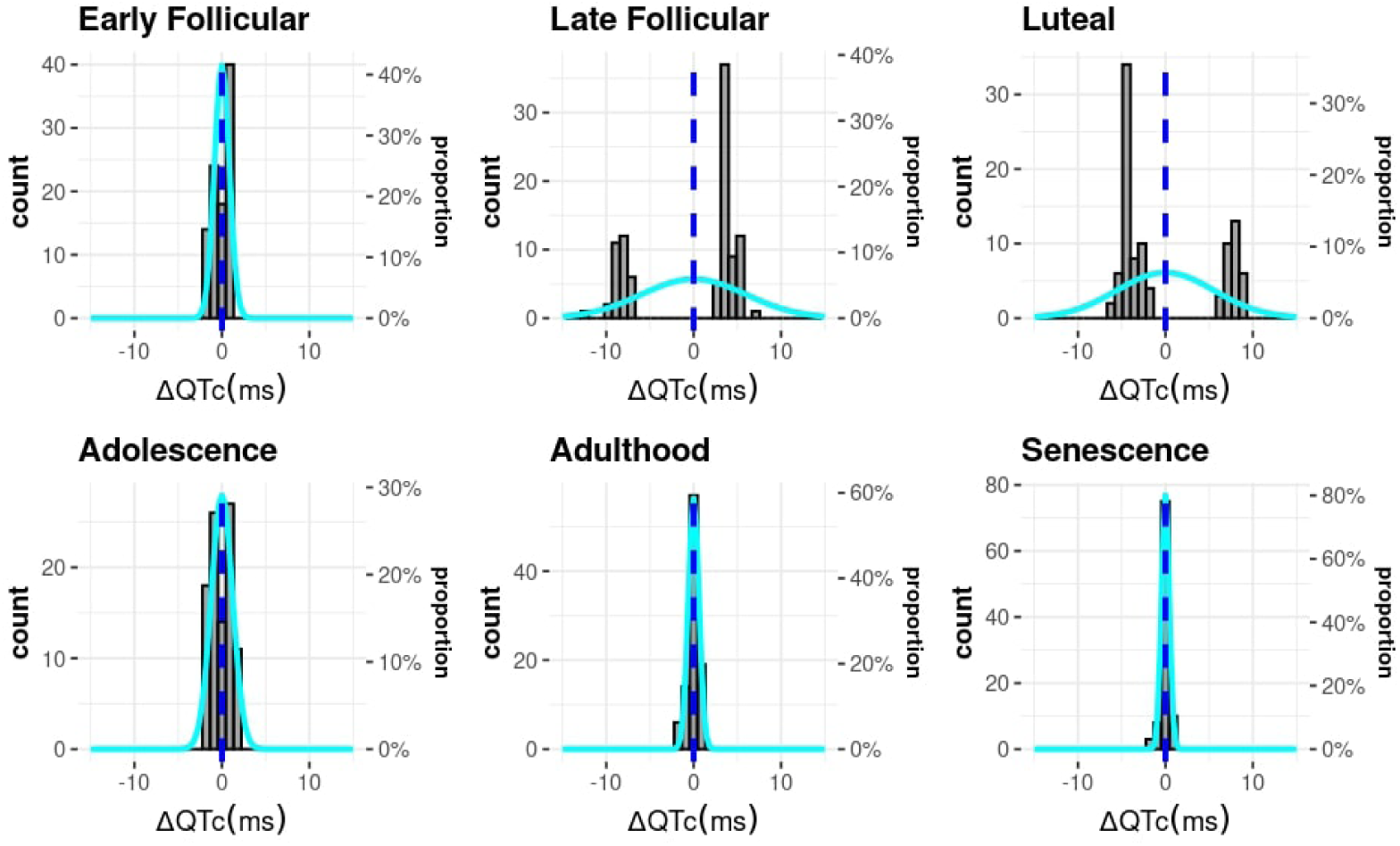
Histograms showing the distribution of ΔQTc of the computational population under placebo effect according to each hormonal stage.

**Figure A.9:**
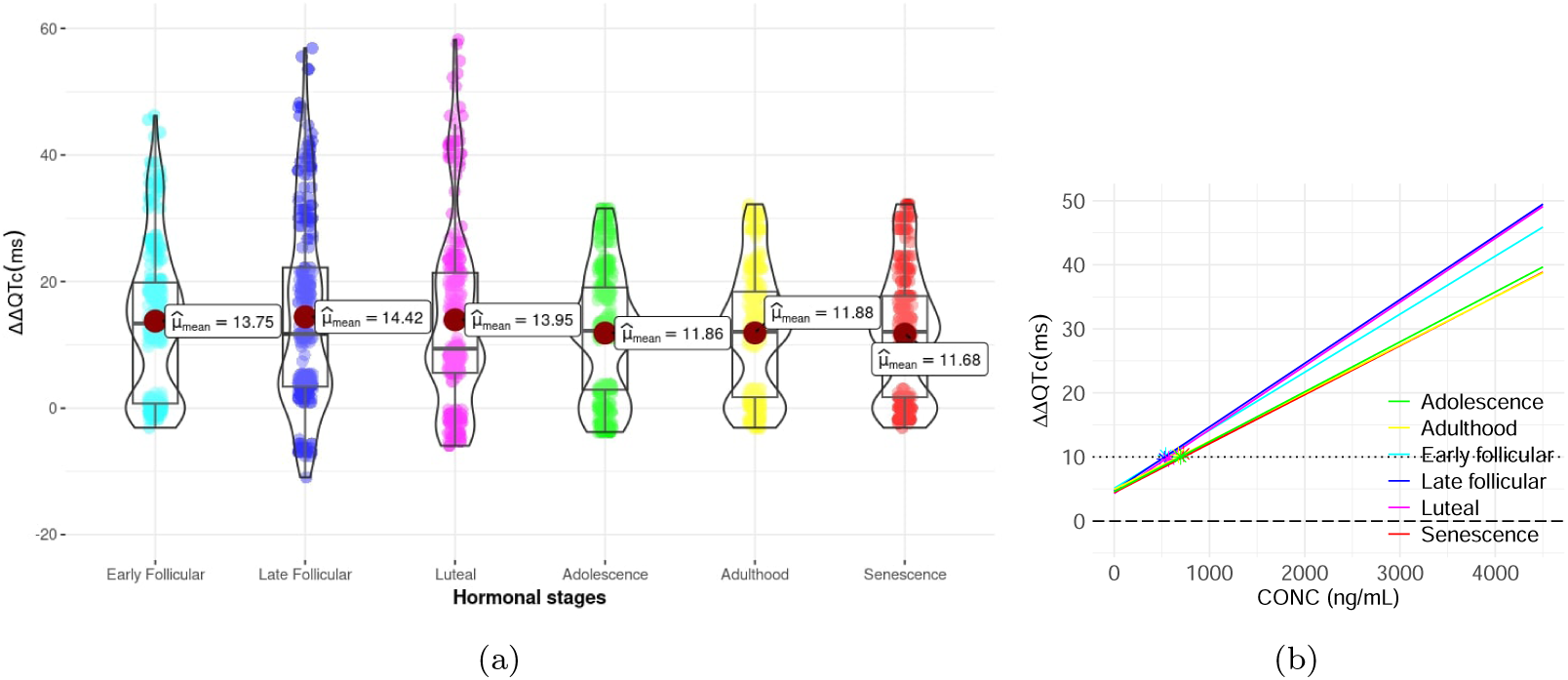
(a) Combination of box and violin plots along with jittered data points of ΔΔQTc in the computational trial under moxifloxacin effect according to each hormonal stage. (b) C-QTc effect models showing the predicted effect of moxifloxacin on ΔΔQTc according to each hormonal stage in the computational trial. Each regression line includes markers to indicate their predicted critical concentration (at the 10 ms threshold).

**Figure A.10:**
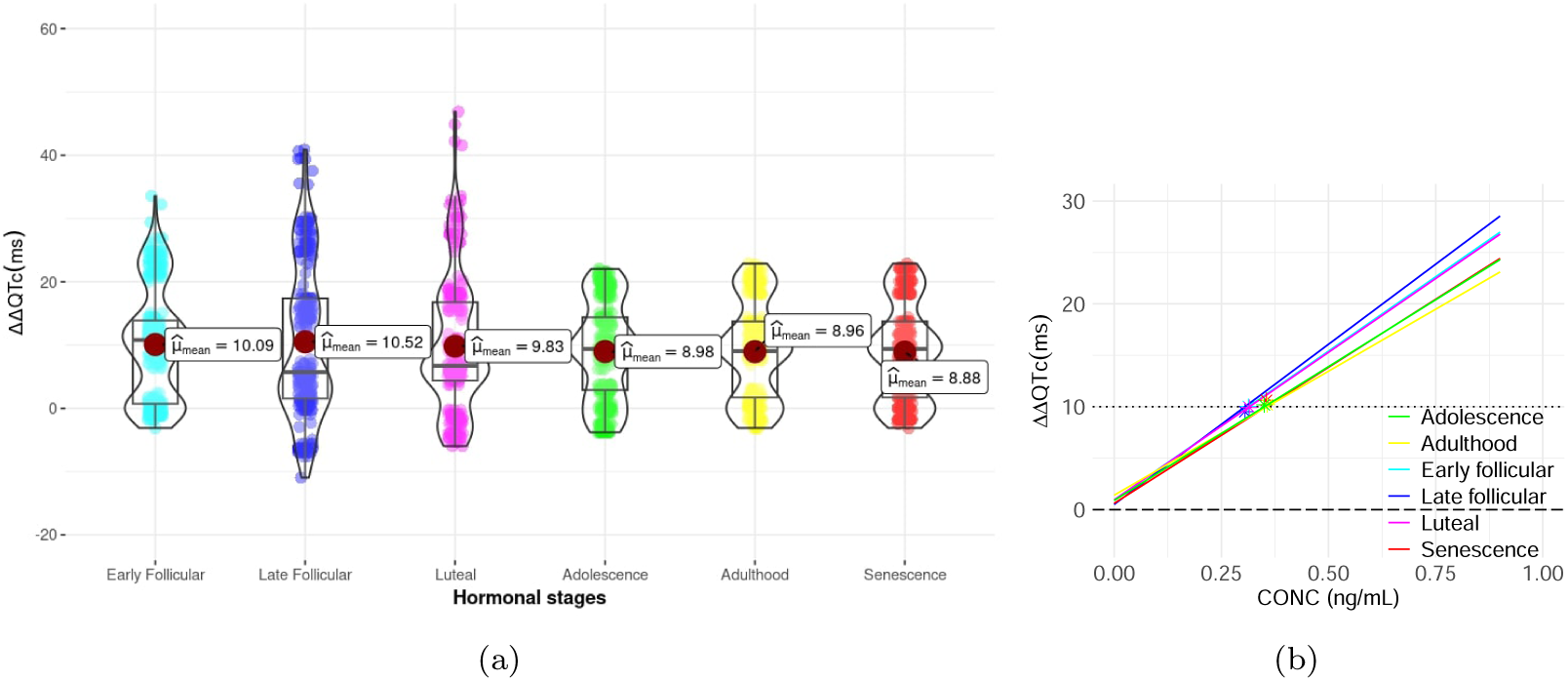
(a) Combination of box and violin plots along with jittered data points of ΔΔQTc in the computational trial under dofetilide effect according to each hormonal stage. (b) C-QTc effect models showing the predicted effect of dofetilide on ΔΔQTc according to each hormonal stage in the computational trial. Each regression line includes markers to indicate their predicted critical concentration (at the 10 ms threshold).

**Figure A.11:**
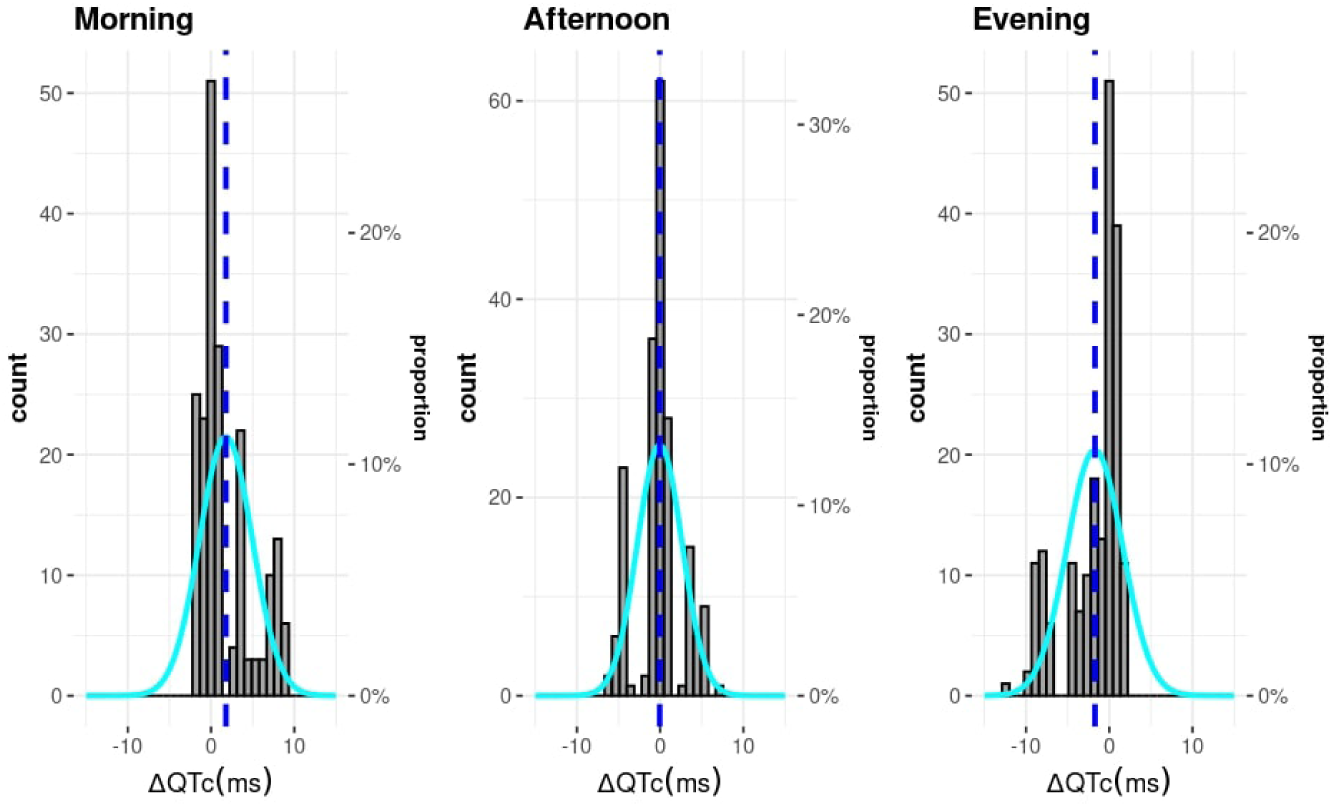
Histograms showing the distribution of ΔQTc of the computational population under placebo effect at different daily time points.

**Figure A.12:**
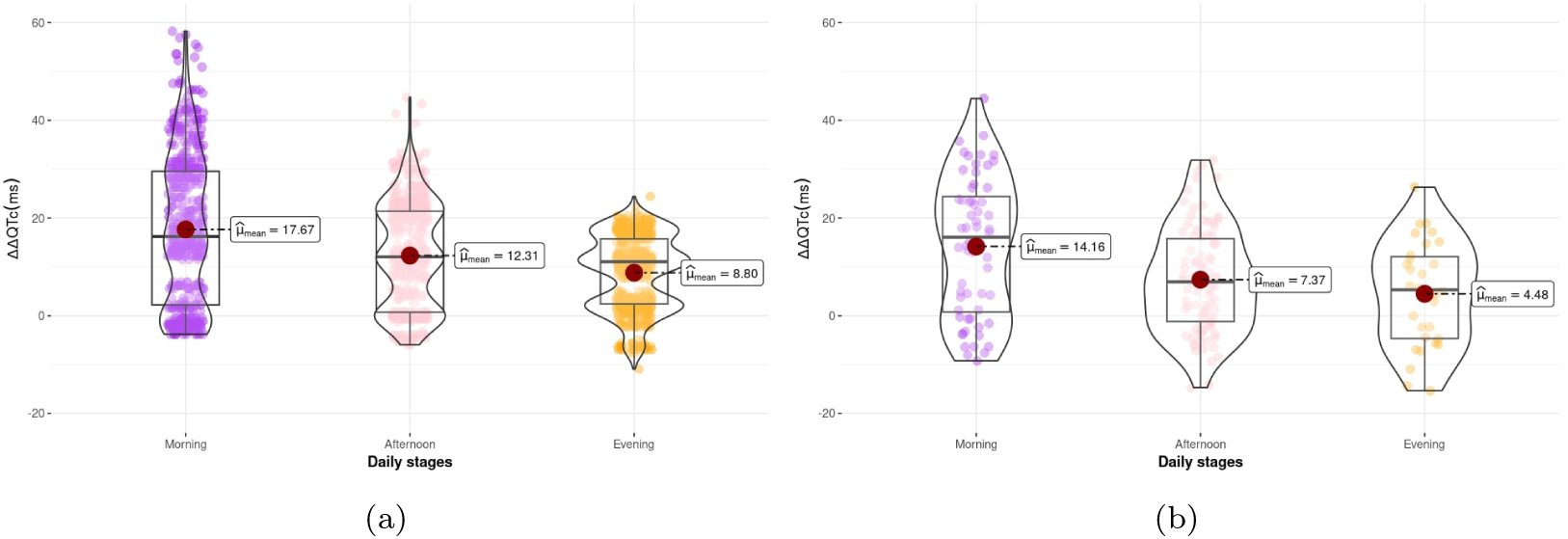
Combination of box and violin plots along with jittered data points of ΔΔQTc in the computational trial (a) and clinical trial (b) after the administration of moxifloxacin at different daily time points.

**Figure A.13:**
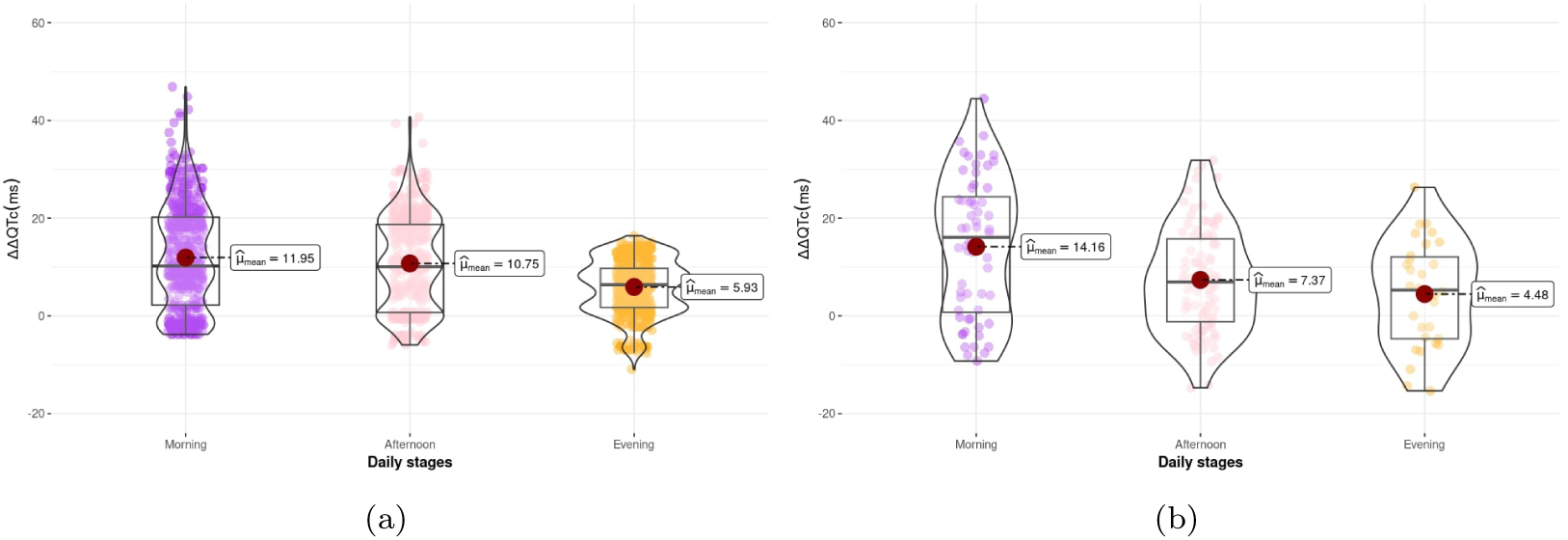
Combination of box and violin plots along with jittered data points of ΔΔQTc in the computational trial (a) and clinical trial (b) after the administration of dofetilide at different daily time points.

**Figure A.14:**
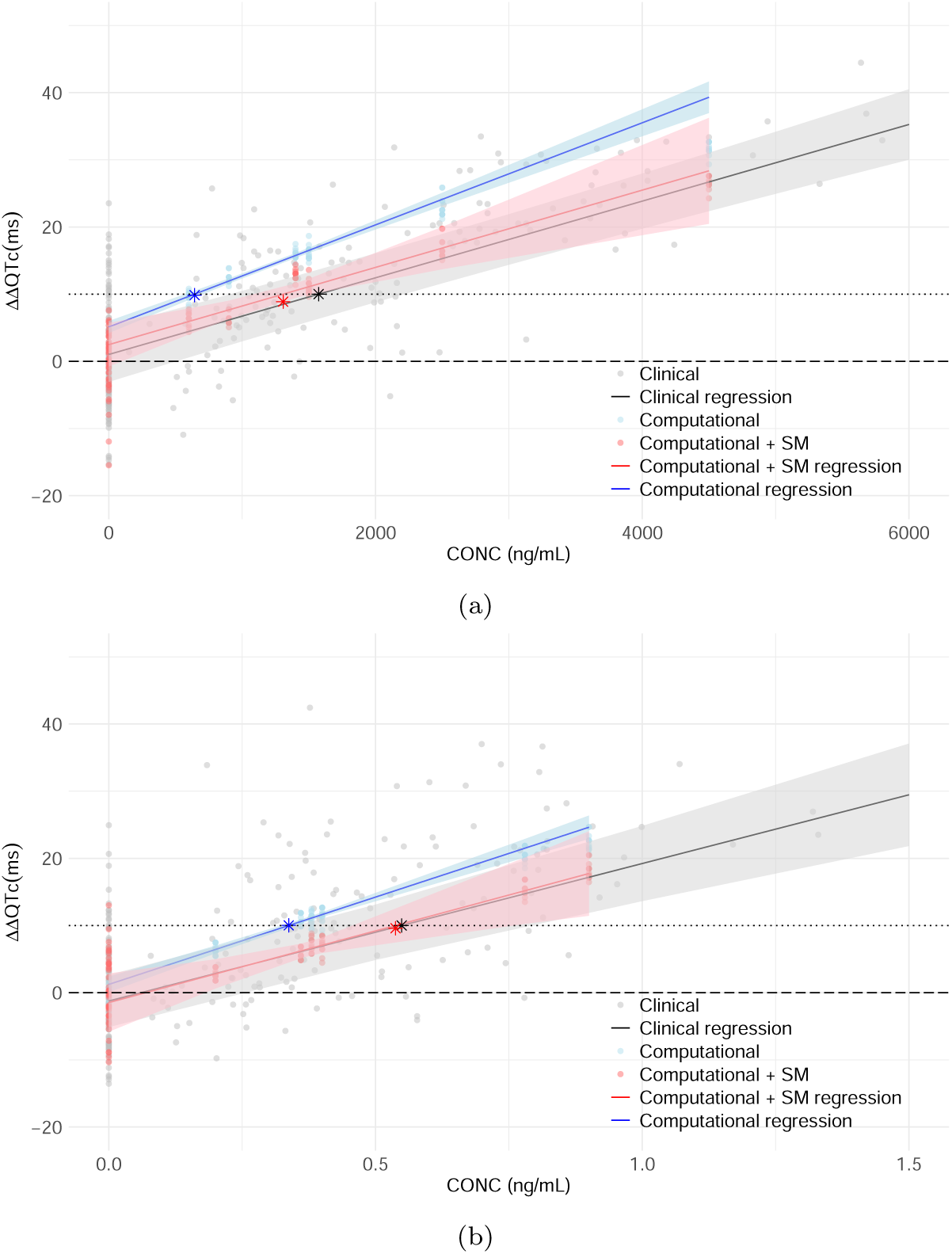
C-QTc effect models showing the predicted effect of moxifloxacin (a) and dofetilide (b) on ΔΔQTc of the population-matched SM-enhanced computational, computational and clinical trials. Each regression line includes markers to indicate their predicted critical concentration (at the 10 ms threshold).

## Footnotes

1 https://www.beta-cae.com/ansa.htm

2 https://go.drugbank.com/drugs/DB00218

3 https://go.drugbank.com/drugs/DB00204

4 https://posit.co/download/rstudio-desktop/

5 https://www.python.org/

6 https://doc.vega.izum.si/

